# Molecular analysis suggests oligoclonality and metastasis of endometriosis lesions across anatomically defined subtypes

**DOI:** 10.1101/2021.04.12.21255355

**Authors:** Teresa H. Praetorius, Anna Leonova, Vivian Lac, Janine Senz, Basile Tessier-Cloutier, Tayyebeh M. Nazeran, Martin Köbel, Marcel Grube, Bernhard Kraemer, Paul J. Yong, Stefan Kommoss, Michael S. Anglesio

## Abstract

Endometriosis symptoms are heterogeneous with controversy on whether it constitutes a single disease or multiple distinct types. Our previous work found recurrent somatic cancer-driver alterations in endometriosis; however, these have not been found ubiquitously. A handful of cases spread across studies also suggest mutations might be shared (clonal) between lesions of the same type. As current classification systems correlate poorly with symptoms or outcomes, somatic genomics may improve the current system. Here, we investigate heterogeneity of somatic cancer-driver mutations within patients and across endometriosis types. We examined anatomically distinct types of endometriosis (ovarian, deep infiltrating, and superficial endometriosis) in 27 individual patients all of whom had at least two types of endometriosis. Specimens were analyzed using high-sensitivity targeted sequencing with orthogonal validation from droplet digital PCR and mutation-surrogate immunohistochemistry. Results found 13/27 patients had informative somatic driver mutation in endometriosis, 9/13 had identical mutations across distinct lesions. Endometriomas tended to have a higher mutational complexity, with functionally redundant driver mutations in same gene and within the same lesions.

Our data are consistent with clonality across endometriosis lesions regardless of subtype. Further the finding of redundancy in mutations with the same gene and lesions is also consistent with endometriosis representing an oligoclonal disease with dissemination likely to consist of multiple epithelial clones travelling together. This suggests the current anatomically defined classification of endometriosis does not fully recognize the etiology of the disease. A novel classification should take into account genomic and other molecular features. These findings could further contribute to development of a more personalized endometriosis diagnosis and care.

## Introduction

Endometriosis is a chronic estrogen-dependent inflammatory disease defined by the presence of endometrial epithelial glands and stroma outside the uterine lumen and a small but significant risk of malignant transformation (1-3). It is estimated to affect up to 10% of biological females of reproductive age and can lead to pelvic pain, dysmenorrhea, dyspareunia, and infertility as well as severely affect quality of life and productivity (2, 4-6). Three major anatomically described types of endometriosis are recognized: superficial peritoneal endometriosis (EM), deep infiltrating endometriosis (DIE), and ovarian endometriomas (OMA) (2, 7). Current classification systems such as the revised American Society for Reproductive Medicine (rASRM) scoring system and the ENZIAN classification for DIE are useful in documenting surgical findings in a standardized manner. However, they poorly correlate with the severity of symptoms and fail to provide a prognostic tool concerning the treatment outcome for pain or infertility (7-9). Likewise, current classification and staging of endometriosis do not include any information about the molecular features or microenvironment of these lesions.

Recent studies have shown that multiple forms of endometriosis harbor somatic cancer-driver alterations including recurrent activating changes in *KRAS, PIK3CA, ARID1A* and others (10-12). It appears the malignant potential for these lesions remains low, despite the presence of recurrent cancer-driver mutations (13-15). However, the contribution of these alterations to the pathobiology of endometriosis remains unclear. Somatic alterations may be useful targets for therapeutic intervention or tracked to study etiology and disease dissemination. The concept that endometriosis disseminates is not novel. Endometriosis frequently presents with multiple anatomical sites affected by lesions (16, 17); recent findings confirm a high rate of coexistence between OMA, EM, and DIE where only 2.3% of study had isolated OMA (16). Along with current etiology largely attributing an origin to endometrial tissues, all endometriosis lesions may well have disseminated from a eutopic point of origin. This highlights the importance of studying the mechanisms of clonal dissemination and metastasis of endometriosis (12, 17, 18). Despite this, there has been little research presenting objective evidence in support of widespread clonal dissemination of endometriosis tissues. Finally, clinical presentation of endometriosis is heterogeneous and it is controversial whether endometriosis constitutes one disease entity or whether independent types with different underlying pathogenesis exist (19).

Here we explored the potential clonal relationship of endometriosis in 27 patients with multiple anatomically separated lesions, each having at least two distinct types of endometriosis. Our objective was assessment of clonality at the level of anatomically described endometriosis types to address whether mutations are frequently shared between lesions and/or between lesion types. If they are not, this would suggest each anatomically defined type represents a unique disease. If they are, this would support plasticity between types and/or that our understanding of endometriosis types is currently insufficient.

## Materials and Methods

### Experimental subject details

Formalin-fixed and paraffin-embedded (FFPE) archival tissues from 27 patients from the Tübingen University Hospital, Germany were included. Inclusion criteria were confirmation of histopathological diagnosis of endometriosis, presentation of two or more types of endometriosis (DIE, and/or EM, and/or OMA) in distinct anatomical locations, and availability of lesions estimated to be of sufficient size for needle macrodissection and yield of DNA for panel sequencing and/or droplet digital PCR, as well as sectioning for immunohistochemistry (IHC). Patients with history of, or co-existing, malignancies were excluded. Clinical diagnosis of endometriosis type was extracted from patient charts and specimens were pathology-reviewed (by Pathologists BTC and TMN) to ensure presence of endometriosis. Experiments were done at the University of British Columbia and the University of Calgary. The project was conducted in compliance with the Canadian Tri-Council Policy Statement on Ethical Conduct for Research Involving Humans (TCPS2, 2018), effort to obtain written informed consent was exercised for all patients. Specimen from non-contactable patients (lost contact/deceased) treated more than 5 years before the start of the study were included under institutionally approved waiver of consent (Tübingen University Hospital Research Ethics Board). All institutions approved use of materials and associated clinical data through local research ethics boards.

### Sample processing and DNA extraction

FFPE specimens were sectioned onto glass slides at 5-8um, stained with dilute hematoxylin and eosin, and manually enriched for endometriosis glands and stroma by needle macrodissection as described previously (11, 20). DNA was extracted using the ARCTURUS® PicoPure® DNA Extraction Kit (ThermoFisher Scientific, USA) and quantitated using the Qubit 2.0 Fluorometer (ThermoFisher Scientific, USA).

### Targeted sequencing

DNA (45-75ng) was sequenced using a proprietary hypersensitive cancer hotspot sequencing panel: FIND IT™, version 3.4 (Canexia Health, Canada) (10, 11). This assay includes hotspots from 33 genes (11, 20) (Table S1). Mutations were considered “true”, if they were genuine hotspot mutations targeted by the FIND IT assay and previously reported in the Catalogue of Somatic Mutations in Cancer (COSMIC) (21), as well as prior observations with validation (10, 11), including a probability score >0.8 and a variant allele frequency (VAF) >0.8% (Table S2A).

### Validation via droplet digital PCR

Somatic mutations identified by targeted sequencing were orthogonally validated through droplet digital polymerase-chain-reaction (ddPCR) assays or IHC (for *TP53* and *PTEN*; see below). In addition, alterations were tested by ddPCR in all available lesions, from a given patient, if they were observed in any one lesion from that given patient in panel-based sequencing. This was done even if panel testing data on other lesions was available, and additionally included any specimens that were omitted from panel testing due to low DNA yield. Using previously established methods (11, 20), extracted DNA was pre-amplified for targets over 10 cycles then diluted before assembling the ddPCR assay. Droplets were generated using the QX200 Droplet Generator (Bio-Rad Laboratories, USA), amplified by thermal cycling, and quantified using the QX2000 Droplet Reader (Bio-Rad Laboratories, USA). As above, alterations were considered “true” if ddPCR droplet counts exceeded the average of the negative control specimens plus 3 times the standard deviation of negative controls the relevant assay. See Table S2A for full listing of results and Table S3 for primer/probe details.

### ARID1A, PTEN and p53 Immunohistochemistry

IHC assays for ARID1A (22, 23), p53 (24, 25) and PTEN (11, 20) (Table S2B, S3) were used as surrogates for somatic alterations following established standards for staining and scoring. In the case of PTEN and p53 we considered loss by IHC (see below) as sufficient for orthogonal validation of somatic mutation found in the mutation panel (none validated; Table S2) or discovery of mutation not covered in the mutation panel. ARID1A was not in our mutation panel but loss was considered as “true” for discovery of somatic mutations.

Scoring was performed as follows: ARID1A loss/mutation, if nuclear staining was absent in endometriosis epithelium cells and internal control (stroma) was intact (22, 23). Mutant p53 (p53abn) if high intensity positive staining was observed in 10 or more adjacent cells in the epithelial cyst wall of endometriosis while maintaining a normal type pattern in surrounding tissue (24, 25). PTEN loss if cytoplasmic and nuclear staining was absent in endometriosis epithelium cells and internal control (stroma) was intact (11, 20). Slides were scored by pathologists TMN, and/or BTC, and/or MK.

ARID1A was stained on Dako Omnis automated immunostainer (Agilent Technologies, USA) or Ventana BenchMark Ultra autostainer (Ventana Medical Systems, USA) the ARID1A rabbit polyclonal antibody, HPA005456 (Sigma-Aldrich). PTEN was stained on Ventana Discovery Ultra (Ventana Medical Systems, USA) using the rabbit monoclonal antibody, 138G6 (Cell Signaling, USA). p53 was stained on Dako Omnis (Agilent Technologies, USA) using the p53 mouse monoclonal antibody DO-7 (GA61661-2; Agilent Technologies, USA).

### Statistics

Student’s t-test was performed to compare the affected genes and lesion types. However, given our limited sample size, they remained non-significant.

## Results

We examined 73 endometriosis lesions from 27 patients with a mean age of 34.9 years (23-45 years, 60% (16/27) of patients were diagnosed with stage IV endometriosis (Table 1; Table S2; Figure S1). 53 lesions were subjected to panel-based sequencing with validation of selected alteration by ddPCR, 6 additional samples included in ddPCR validation only. We relied primarily on mutation data to establish clonality (Table S4), but also included supportive mutation surrogate IHC data for ARID1A, PTEN, and p53. Tumor suppressors ARID1A and PTEN are frequently altered in endometriosis associated ovarian cancer and have been reported to be somatically altered at varying frequencies in endometriosis (11, 13, 23, 26, 27). Somatic alteration of p53 is less commonly reported in endometriosis and endometriosis associated carcinomas (28-30). While our mutation panel has good coverage of the *TP53* gene the p53 IHC assay provided validation as well as indication of mutations not covered in the panel assay. In total, we IHC assayed lesions for PTEN (66 interpretable), ARID1A (66 interpretable) and p53 (47 interpretable). Including mutation and IHC data a total of 27/59 (45.8%) lesions from 13/27 (48%) cases had identifiable somatic cancer-driver mutations.

**Table 1:**
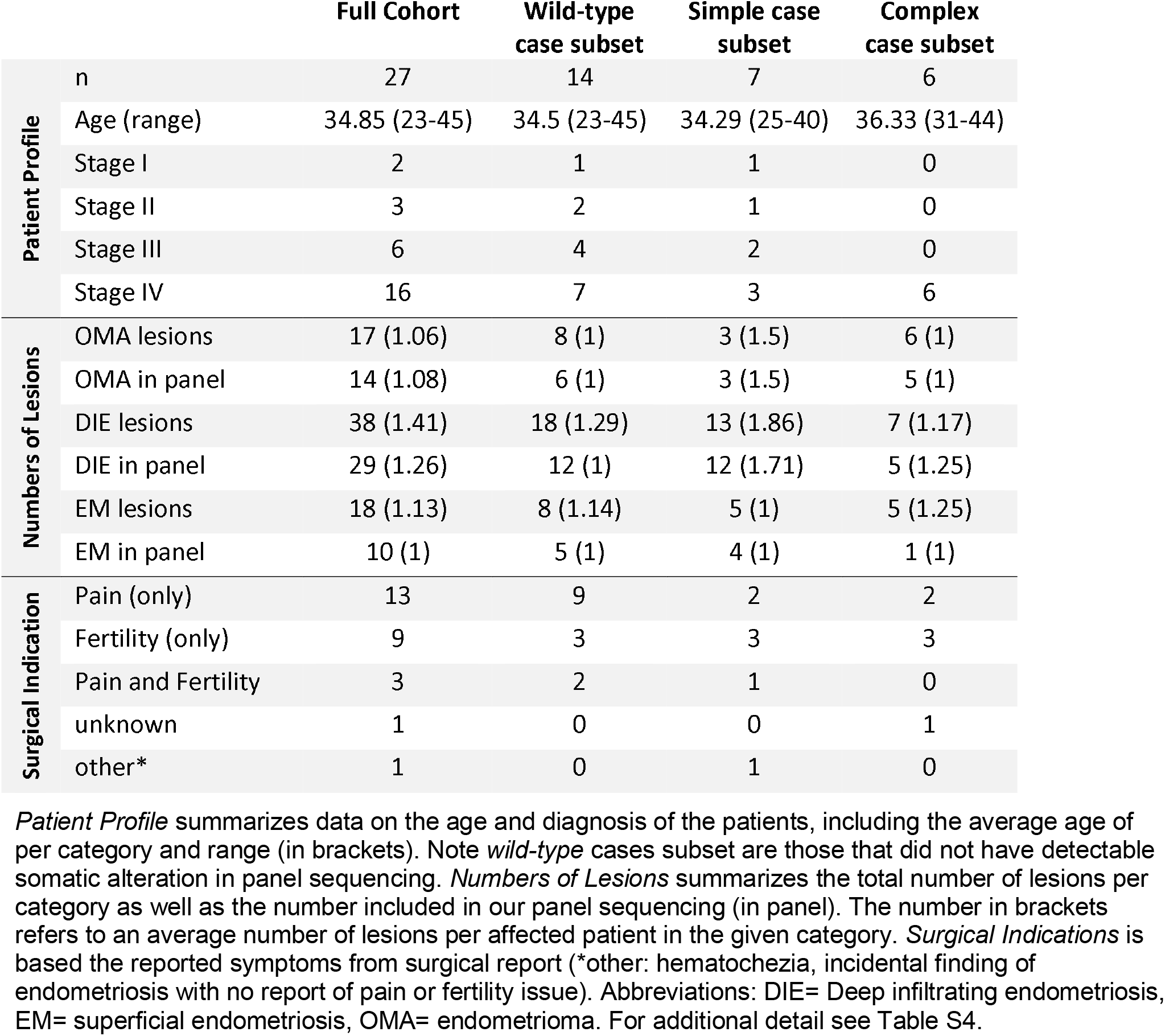
Clinical characteristics of endometriosis cases categorized by mutation profile.

|  |  | Full Cohort | Wild-type<br>case subset | Simple case<br>subset | Complex<br>case subset |
| --- | --- | --- | --- | --- | --- |
| Patient Profile | n | 27 | 14 | 7 | 6 |
|  | Age (range) | 34.85 (23-45) | 34.5 (23-45) | 34.29 (25-40) | 36.33 (31-44) |
|  | Stage I | 2 | 1 | 1 | 0 |
|  | Stage II | 3 | 2 | 1 | 0 |
|  | Stage III | 6 | 4 | 2 | 0 |
|  | Stage IV | 16 | 7 | 3 | 6 |
| Numbers of Lesions | OMA lesions | 17 (1.06) | 8 (1) | 3 (1.5) | 6 (1) |
|  | OMA in panel | 14 (1.08) | 6 (1) | 3 (1.5) | 5 (1) |
|  | DIE lesions | 38 (1.41) | 18 (1.29) | 13 (1.86) | 7 (1.17) |
|  | DIE in panel | 29 (1.26) | 12 (1) | 12 (1.71) | 5 (1.25) |
|  | EM lesions | 18 (1.13) | 8 (1.14) | 5 (1) | 5 (1.25) |
|  | EM in panel | 10 (1) | 5 (1) | 4 (1) | 1 (1) |
| Surgical Indication | Pain (only) | 13 | 9 | 2 | 2 |
|  | Fertility (only) | 9 | 3 | 3 | 3 |
|  | Pain and Fertility | 3 | 2 | 1 | 0 |
|  | unknown | 1 | 0 | 0 | 1 |
|  | other* | 1 | 0 | 1 | 0 |

Amongst the panel screened lesions, nearly half of the tested cases had at least one mutation. *PIK3CA* alterations were the most common (27 hotspot mutations, affecting 12/53 lesions in 6 cases), followed by *KRAS* (16 hotspot mutations, affecting 15 lesions in 6 cases) and *CTNNB1* (10 hotspot mutations, affecting 7/53 lesions in 4 cases; Figure 1). In contrast to finding more *PIK3CA* alterations than any others, more lesions were affected by *KRAS* alterations. This trend was the same regardless of lesion type (Figure 1D) or if summarizing by fraction of affected cases (Figure 1E). Further, OMA tended to have a higher proportion of lesions affected by somatic cancer-driver alterations and subsequently higher mutation load than other lesion types (Figure 1, Table S2).

**Figure 1:**
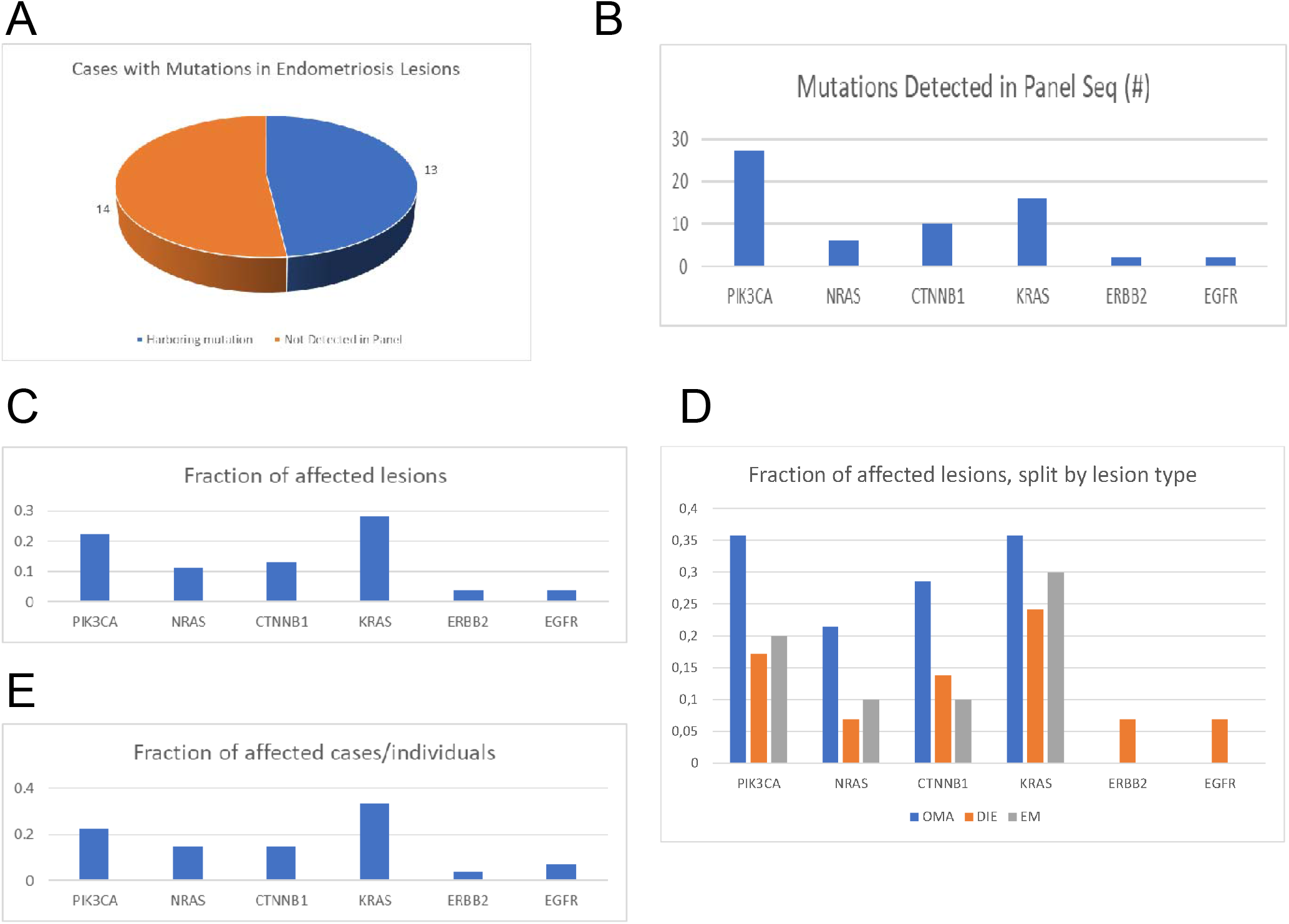
Fraction of cohort affected by somatic hotspot mutations detected in panel sequencing. (A) Overall split of cases with at least one lesions having at least one somatic cancer-driver alteration. (B) Numbers of detected somatic mutations, split by affected gene, across our entire cohort. (C) Fraction of mutation affected lesions, split by mutation type. (D) Fraction of mutation affected lesions, split by lesion type. (E) Fraction of mutation affected lesions, summarized by fraction of affected cases.

Alterations were less frequently observed in *NRAS* (6 mutations, 6/53 lesions, 4 cases), *ERBB2* (2 mutations, 2/53 lesions, 1 case) and *EGFR* (2 mutations, 2/53 lesions, 2 cases). Although our cohort had insufficient numbers to support strong associations, we noted many lesions, predominantly amongst OMA, contained multiple mutations in the same genes.

No samples had identifiable p53-abnormal staining pattern (0/47 lesions), PTEN loss was observed in 5 cases (8/66 lesions), and ARID1A loss was observed in 1 case (2/66 lesions). Immunohistochemical data is inconclusive with respect to clonality, and similar abnormal patterns were considered in support of mutation findings. Our mutation data suggested a single *TP53* alteration but no abnormal p53 IHC pattern was observed. As IHC for p53 is accepted to be a surrogate for mutation status we considered this as a false positive (case 22; Table S2A) (24, 25, 31). We observed no *PTEN* mutations in sequencing data despite evidence of loss in PTEN protein expression by IHC in 8/66 lesions (Table S2B), however only a few *PTEN* hotspots are covered in panel sequencing (Table S1). In case 2, with PTEN loss in multiple lesions, other somatic (clonal) point mutations were shared between the PTEN-loss affected lesions (Table S2A). Case 3 showed loss of ARID1A protein consistent with loss-of function somatic mutation (22) in both an EM and OMA samples (Figure 2). Unfortunately, insufficient tissue was available to validate any mutation in the EM sample from Case 3, however, the *PIK3CA* (p.Met1043Ile) and *KRAS* alterations were shared across all other lesions from this case. Altogether, we observed clonality between at least two endometriotic lesions in 8 out of 13 informative cases (Figure S2; Table S2; Table S4).

**Figure 2:**
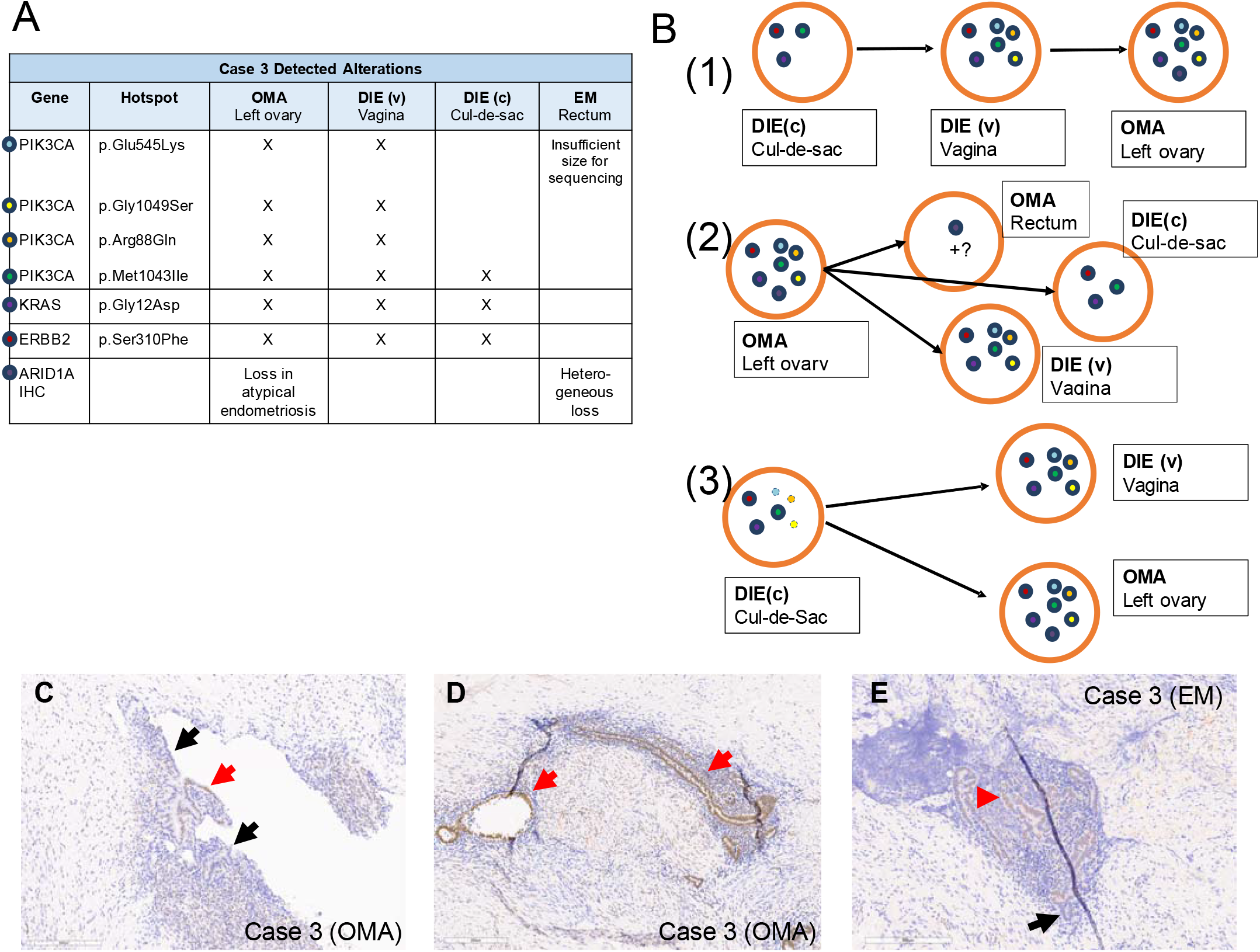
Clonal relationships in Case 3 patient presenting with DIE of the vagina, DIE of the cul-de-sac, an OMA of the left ovary, and EM of the rectum. (A) Detected somatic alteration from sequencing and mutation surrogate IHC suggest identical/clonal alteration in *KRAS, ERBB2*, and *PIK3CA* (p.Met1043Ile) between OMA and two anatomically distinct lesions of DIE. Additional *PIK3CA* alterations are also visible in the OMA and vaginal DIE. Heterogeneous loss of ARID1A is observed in only the OMA and EM (insufficient material was available for mutation testing in EM). (B) Three hypothetical dissemination models explaining the mutational pattern: (1) top panel illustrates a linear pattern wherein clones from the cul-de-sac travel to the vagina and acquired additional alterations in *PIK3CA*. Subsequent transfer of all clones to the ovary where subclonal-ARID1A loss occurs. It may further be speculated that the EM resulted from a transfer of clones between the vaginal DIE and OMA (as the initial site for ARID1A loss) or subsequent spread after clones established on the ovary. Given the lack of mutational data on the EM lesion it may also be entirely independent. (2) middle panel illustrates an example where a complex set of clones exist at the ovary and only a subset of these break-off and colonize vaginal, cul-de-sac and EM/rectal sites. Finally, (3) Lower panel illustrates parallel dissemination from the cul-de-sac lesion to both the vaginal and ovarian sites. Herein we may consider sub-threshold signal from ddPCR of p.Arg88Gln, p.Glu545Lys and p.Gly1049Ser PIK3CA alteration (Table S2) as weak evidence of emerging/undetectable clones that subsequently expand post-transfer of cells to both vagina and ovary. In this model, the OMA acquires loss-of function ARID1A alterations. Additional possibilities may also explain the mutational patterns. (C-D-E) photomicrographs showing ARID1A IHC results including regions of loss (surrogate for loss-of function mutation; black arrows) and normal (retained nuclear staining; red arrows) in endometriosis epithelium.

For ease in presenting results details we have divided informative cases (n = 13) into two categories (Table 1; Table S4). *Simple cases* had only one or two altered genes and/or one or two informative lesions. *Complex cases* all had alterations across lesions and at least one lesions with functionally redundant alterations (i.e. equivalent activating change resulting from different nucleotide and/or amino acid substitution in the same gene).

### Simple cases

Seven cases were defined as simple (Table 1). In 3 of these we identified and validated shared mutations across multiple lesions and types (Figure S2; Table S4). Case 6 shared mutations in *PIK3CA* between EM and DIE. The other two cases (19 and 21) had mutations in *KRAS*. Case 19 shared a p.Gly12Ser mutation between an EM and a DIE. Case 21 shared a p.Gly12Asp between an OMA and three different DIE lesions.

The 4 remaining cases had somatic alteration that were not shared across lesions within individuals, thus were not informative with respect to clonality (Figure S2). Of these, 3 cases (20, 22, 24), had mutations observed in only a single lesion (*KRAS* or *EGFR*) while 1 (Case 9) had alterations in KRAS and PIK3CA but none shared between lesions. Case 9 did have two (redundant) KRAS alterations in DIE but we retained this case in our “simple” category as no alterations were shared between lesions.

### Complex Cases

All six cases in the complex category were stage IV and showed evidence of a clonal relationships between lesions (Table 1, Table S4). In each case at least one lesion showed multiple functionally redundant hotspot alterations (Figure S2). We observed identical somatic alterations shared between EM and DIE lesions in one case (case 13); between DIE and OMA lesions in 4 cases (cases 1, 2, 3, 8; Figure 2); and between all three types in one case (case 4; Figure 3).

**Figure 3:**
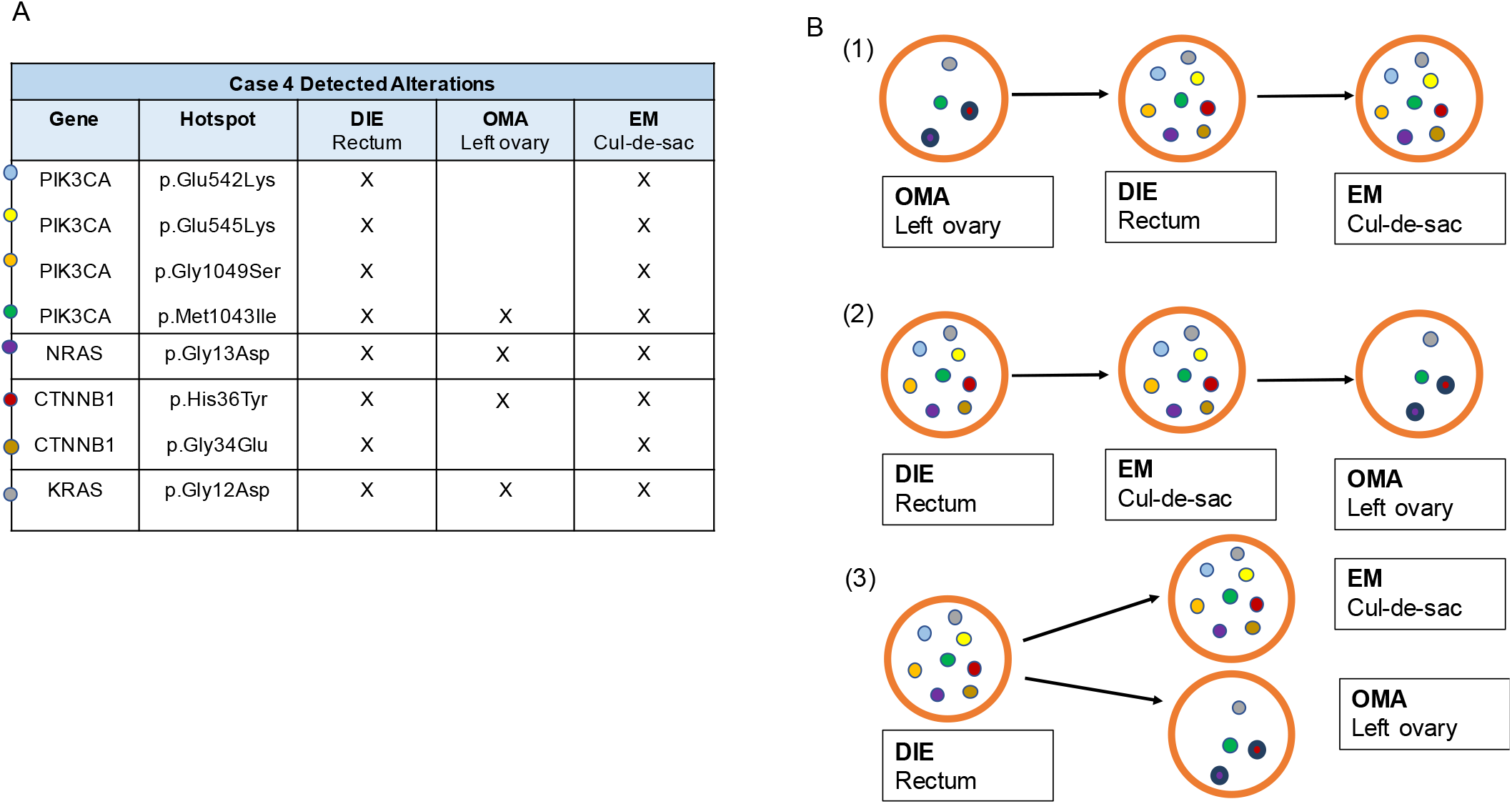
Clonal relationships in Case 4 patient presenting with DIE of the rectum, EM of the cul-de-sac and an OMA on the left ovary. (A) Detected somatic alteration from sequencing suggest all variants are shared between DIE and EM lesions. In contrast the ovarian lesion shares only 1 *PIK3CA* and 1 *KRAS* alteration with the other two and has no (detected) unique changes. (B) Three possible dissemination models explaining the mutational pattern: (1) top panel illustrates a linear model wherein clones from the ovary travel to the rectum, expand and acquire additional alteration. Cells from the DIE containing all clones then seed the EM on the Cul-de-sac. (2) middle panel illustrates another linear model of dissemination wherein a complex clonal population at the Rectum seed the cul-de-sac. Subsequently only a subset of these clone travel to and populate the ovary. Finally, (3) illustrates a differing model of spread wherein a complex clonal population at the Rectum seed the cul-de-sac with all clones. Only a subset with a single *PIK3CA* and *KRAS* altered clone break-off, or are capable of, colonizing the ovary.

Multiple co-existing activating mutations (functional redundancy) were most common with *PIK3CA*: seen in 12 lesions across 5 cases. In 4/5 cases with *PIK3CA* mutations we observed co-existing alterations in *CTNNB1*, and in all of these at least one was shared across multiple sites/types within the affected patient.

*KRAS* alterations were seen in 5 of 6 *complex cases*, only case 1 in this category showed multiple redundant activating changes (p.Gly12Val in DIE, p.Gly12Ser in both DIE and OMA). As *PIK3CA, CTNNB1* and *KRAS* alteration were all relatively common we saw co-existence between all of these in 3/6 *complex cases*.

*NRAS* alterations were detected in 4 of 6 *complex cases*. In 3 cases (Cases 1, 2, 8) an *NRAS* alteration was shared between OMA and DIE. In case 4 we detected one *NRAS* alteration shared between OMA, DIE, and EM. None of the cases showed redundant alterations of *NRAS* itself but they did overlap with *KRAS* mutations.

*ERBB2* alterations were detected in 2 of 6 *complex cases*. In case 3 this was shared between an OMA and two DIE lesions (case 3) and co-occurred with a p.Gly12Asp KRAS mutation at all three sites. In case 13, the ERBB2 mutation was seen only in EM but co-occurred with an EGFR mutation. The same EGFR mutation was also present in DIE from case 13.

## Discussion

We examined a total of 73 lesions from 27 individuals affected by endometriosis and focused on within patient heterogeneity – a largely overlooked feature amongst current genomic studies on endometriosis. Cases were specifically selected to have multiple anatomical sites affected as well as multiple “types” of endometriosis: EM, DIE and OMA. Using a high-sensitivity, and error-correcting, sequencing technology followed by validation and additional screening with ddPCR we identified somatic cancer driver alterations in lesions from 13 patients. A total of 27 lesions had at least one driver alteration, 7 lesions had more than 5.

We found evidence of identical alterations across lesions in 10 individuals, all of which spanned more than one type of endometriosis. The relatively commonplace finding of identical mutations across lesions suggests that at least a subset are clonal: sharing a common ancestor within a given patient. According to our present data set, the order/directionality of dissemination cannot be determined.

In 9 lesions there also appeared to be multiple functionally redundant alterations, which is consistent with individual lesions being oligoclonal (i.e. comprised a small number of clones), and often spread across multiple anatomically distinct lesions. Functionally redundant mutations were seen in *PIK3CA* (activating), *CTNNB1* (stabilizing), *KRAS* (activating), and more generally activating Ras-pathway (e.g. *KRAS, NRAS, ERBB2, EGFR*). Redundant mutation in the same gene were seen in 6 cases, half of which had the redundant alterations in two or more lesions. Intra-lesion Ras-pathway redundancy, including activating somatic alterations in the same gene or different Ras-pathway genes, affected 9 lesions in 5 cases.

Examining the specific prevalence of mutations across lesions we observed more *PIK3CA* mutations than any other. Amongst informative cases, OMAs tended to have more mutations, with OMA lesions from cases 1, 2, 3 having multiple activating *PIK3CA* and/or *CTNNB1* (and others) driving the average mutation burden up. In contrast, DIE appeared to have a wider spectrum of mutations with *EGFR* and *ERBB2* mutations seen only in DIE. Despite these raw numbers, more lesion and more individuals appeared to be affected by at least one Ras-pathway activating alteration, most frequently *KRAS*.

Our study is unique in examination of a large number of co-existing lesions per patient, allowing us to analyze both molecular heterogeneity and potential clonal dissemination/metastasis. Particular strengths include enrichment of endometriosis glands and stroma, the use of proven high-sensitivity and specificity digital sequencing methods, and orthogonal validation for the majority of alterations (13).

Due to application of manual macrodissection over laser capture microdissection we did not isolate (stromal/epithelial) cell types and can only use the variant allele frequency for binary presence or absence assessment of somatic alterations. IHC observations suggested somatic alterations were exclusive to the epithelium, similar to prior reports (10, 11, 32). In addition, we used only a relatively limited panel assay to screen for mutations, albeit studies that have undertaken exome analysis of endometriosis have most frequently reported somatic alterations in genes where our mutation panel has coverage (10, 12, 32, 33). Nonetheless, while we can make suggestions about potential lineage of samples with informative mutations, we are missing considerable data about the un-analyzed genome. In these cases, panel sequencing and subsequent ddPCR often required re-sampling for validation. The fraction of enriched endometriosis cells harboring mutation may change as the specimen is sectioned/sampled, and that it would genuinely yield a fraction of cells below a detection threshold in one sampling and above a threshold in another. Thus, this should be considered as a minimum estimate of clonality. Lesions (within a given patient) that do not share mutations cannot be definitively determined to be of unrelated lineage.

In recent years there has been a shift away from seeing endometriosis as one disease entity. In addition to biochemical variations, considerable efforts are being applied to identify subtype-specific genomic alterations. Our findings validate a modest spectrum of driver alterations, as has been described previously (10, 12, 32, 33), while shared alterations between lesions support a model of metastasis. Our observations allowed us to partially categorize cases based on mutation load and distribution: those without detectable mutations, *simple*, and *complex* cases. It is worthwhile noting that all *complex cases*, with higher mutation burden as well as gene and pathway redundancy, were stage IV while other categories showed a range including low stage cases. Further, gene and pathway redundancy in observed mutations, suggest endometriosis lesions may be comprised of a small number of clones within the epithelium and that metastasis is also oligoclonal: these clones may disperse and travel together.

To make significant impact the next generation of endometriosis-genomic studies must apply high stringency methods, with appropriate enrichment of tissues, orthogonal validation and/or error correcting sequencing technologies, and most critically be coupled to large clinical data registries conforming to accepted standards in phenotyping data (13) - such as those promoted by WERF-EPHect (34, 35). Early studies on somatic genomics of endometriosis have suggested relatively few alterations per lesion (10, 12, 32, 36). However, this work suggests endometriosis may have moderate genomic complexity. Any future study wishing to correlate genomic heterogeneity with the burden of disease (stage) and clinical phenotypic spectrum must take this into account. Accurate assessment of somatic mutation profiles may require a significant fraction of lesions are excised and tested at surgery (rather than ablated without biopsy).

This work has also shown that OMAs had the highest potential for oligoclonality, while DIE lesions were associated with wider range of mutations. Considering previous work in the field (37-40), this is consistent with the concept that distinct microenvironments are important for lesion forming and spreading. The ovarian microenvironment may provide permissive conditions for expansion of multiple clones which could be associated with the higher risk of malignant transformation (41). DIEs might be surrounded by a more restrictive microenvironment conditions resulting in fewer clones and lower malignant potential. As a potential example: redundant *PIK3CA* activating mutation were prevalent in OMAs. Such *PIK3CA* alterations are common in endometriosis-associated cancers, and appear necessary (in combination with ARID1A loss of function) for the generation of a representative clear cell ovarian carcinomas model in mice (42). Thus, OMAs with multiple *PIK3CA* mutant clones may have elevated malignant potential. In contrast, the extraovarian microenvironment restriction on malignant potential has been suggested as a potential mechanism for the generally favorable outcomes observed in low-stage synchronous (yet metastatic) endometrioid ovarian and endometrial carcinomas (43).

## Conclusion

Our results provide conclusive data for two major features of endometriosis: first that the disease has a high potential for molecular heterogeneity as shown by mutation profiles indicating oligoclonality, or at least clonal divergence. Second, endometriosis is capable of metastasis and this is not restricted to current definitions of endometriosis types. These features confirm that the current classification of endometriosis is limited. While our series is not sufficient to suggest which molecular features should be included in a novel classification, we can unequivocally state future work should consider patient-wide endometriosis mutation profiles – not single lesions. This may include ranking of allele frequency, clonality across lesions, mutation burden (or clonal burden), and anatomic types. Intra-lesion spatial heterogeneity (such as in a large OMA or deep nodules) and clonal burden also warrants study especially in relapse/persistent chronic disease and those associated with malignant progression.

## Data Availability

All data is contained within the manuscript and/or associated supplemental information.

## Acknowledgements

The authors thank all the study participants who contributed to this study. We further recognize the invaluable contributions of Prof. Dr. Sara Y. Brucker for continuous support of international collaborations between the University Hospital Tübingen and the University of British Columbia, as well as PD Dr. Annette Staebler and the staff of the Institute of Pathology, University Hospital Tübingen for facilitating access to pathology archives.

Funding was provided by Canadian Institutes of Health Research (Early Career Investigator Grant in in Maternal, Reproductive, Child & Youth Health to MS Anglesio). M Köbel received support through the Calgary Laboratory Services research support fund (RS19-609). MS Anglesio is funded through a Michael Smith Foundation for Health Research Scholar Award (18274) and the Janet D. Cottrelle Foundation Scholars program (managed by the BC Cancer Foundation). This project received technical and data management support from Calgary Laboratory Services and the Genetic Pathology Evaluation Centre (GPEC). GPEC receive core support from BC’s Gynecological Cancer Research team (OVCARE), and the VGH+UBC Hospital Foundation. M Köbel was supported by internal research support (RS19-621).

## Supplemental Figure and Table Legends

**Figure S1:** Project workflow. 27 patients were selected all with more than one type and more than one anatomical site of endometriosis. A total of 73 lesions were reviewed for analysis. 53 lesions had sufficient tissue for digital panel sequencing while an additional 6 samples had sufficient yield for only ddPCR of select alterations. A total of 83 somatic alterations were initially detected in sequencing, 72 subjected to ddPCR validation (58 validated). During the ddPCR validation stage additional lesion were tested for alterations observed in panel sequencing resulting in a total of 90 detected alterations in 13 cases across the entire cohort. Immunohistochemistry for PTEN (loss in 11%), ARID1A (loss in 3%), and p53 (no abnormal staining observed) was done in parallel.

**Figure S2:** Per-Case summary of detected alterations across lesions as derived from sequencing, digital droplet PCR (ddPCR), and immunohistochemistry (IHC) in informative cases. Legend: Abd = abdominal, atyp= atypical, DIE= deep infiltrating endometriosis, EM= superficial endometriosis, Het loss= heterogeneous loss, l= left, lig= ligament, pararec=pararectal, OMA= endometrioma, r=right rv= rectovaginal, su= sacrouterine

**Table S1:** Hotspots and Exons analyzed in the FIND IT™ panel version 3.4 (Canexia Health, Canada)

**Table S2A:** Panel Sequencing and ddPCR validation data. Full list of sequence and droplet digital PCR (ddPCR) data with variant allele frequencies (VAFs) and probability scores (PR.)

DIE= deep infiltrating endometriosis, EM= superficial endometriosis, OMA= endometrioma

**Table S2B:** Immunohistochemistry. Full list of IHC results, DIE= deep infiltrating endometriosis, EM= superficial endometriosis, OMA= endometrioma

**Table S3:** Resource table of antibodies, primers, and probes

**Table S4:** Cohort Overview. Each cell shows the number of lesions available and tested from the noted anatomical site/type (number in brackets denote lesions were not tested in panel sequencing and detected only in ddPCR). Blue cell = shared alteration within case/between lesions marked by blue cells in table (cells not in blue were uninformative/did not show evidence of clonality with any other lesion from the given case), Grey lines/cases were entirely uninformative (no mutation detected in panel sequencing from any tested lesion). Abbreviations: DIE= Deep infiltrating endometriosis, EM= superficial endometriosis, OMA= endometrioma

