## Supplemental Figures S1-S2 for "Molecular analysis suggests oligoclonality and metastasis of endometriosis lesions across anatomically defined subtypes"

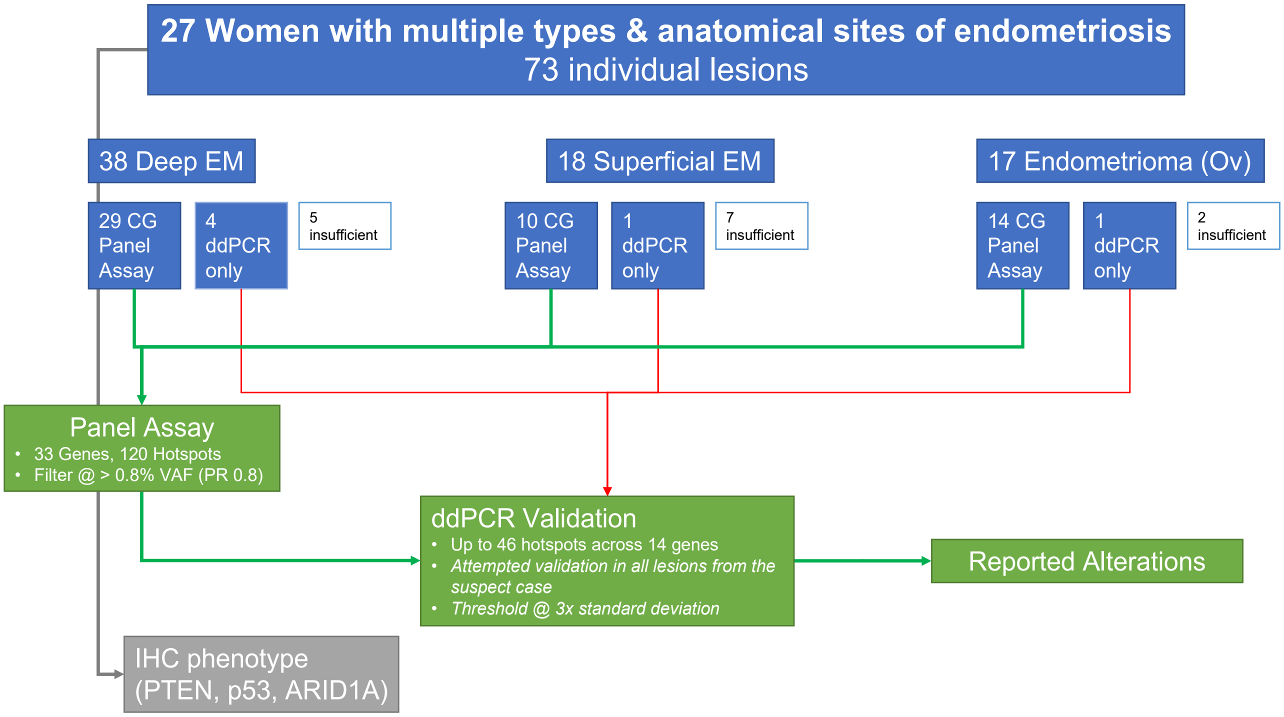


**Figure S1:** Project workflow.

27 patients were selected all with more than one type and more than one anatomical site of endometriosis. A total of 73 lesions were reviewed for analysis. 53 lesions had sufficient tissue for digital panel sequencing while an additional 6 samples had sufficient yield for only ddPCR of select alterations. A total of 83 somatic alterations were initially detected in sequencing, 72 subjected to ddPCR validation (58 validated). During the ddPCR validation stage additional lesion were tested for alterations observed in panel sequencing resulting in a total of 90 detected alterations in 13 cases across the entire cohort. Immunohistochemistry for PTEN (loss in 11%), ARID1A (loss in 3%), and p53 (no abnormal staining observed) was done in parallel.

Figure S2

**Detected Alterations in Complex Cases**

**Case 1 Detected Alterations**

| Gene | Hotspot | OMA R.Ovary | DIE Rectum |
|---|---|---|---|
| PIK3CA | p.Glu542Lys | X | |
| PIK3CA | p.Gly1049Ser | X | X |
| PIK3CA | p.Arg88Gln | X | |
| PIK3CA | p.Met1043Ile | X | X |
| NRAS | p.Gly13Asp | X | X |
| CTNNB1 | p.His36Tyr | X | |
| CTNNB1 | p.Gly34Glu | X | X |
| CTNNB1 | p.Thr41Ile | X | X |
| KRAS | p.Gly12Ser | X | |
| KRAS | p.Gly12Val | | X |

**Case2 Detected Alterations**

| Gene | Hotspot | OMA R.ovary | DIE Rectum |
|---|---|---|---|
| PIK3CA | p.Glu542Lys | X | X |
| PIK3CA | p.Glu545Lys | X | |
| PIK3CA | p.Gly1049Ser | X | |
| PIK3CA | p.Arg88Gln | X | |
| PIK3CA | p.Met1043Ile | X | |
| NRAS | p.Gly13Asp | X | X |
| CTNNB1 | p.His36Tyr | | X |
| CTNNB1 | p.Gly34Glu | X | |
| CTNNB1 | p.Thr41Ile | X | |
| PTEN | IHC | Het loss | Het loss |

**Case 3 Detected Alterations**

| Gene | Hotspot | OMA L.ovary | DIE Vagina | DIE Cul-de-sac |
|---|---|---|---|---|
| PIK3CA | p.Glu545Lys | X | X | |
| PIK3CA | p.Gly1049Ser | X | X | |
| PIK3CA | p.Arg88Gln | X | X | |
| PIK3CA | p.Met1043Ile | X | X | X |
| KRAS | p.Gly12Asp | X | X | X |
| ERBB2 | p.Ser310Phe | X | X | X |
| ARID1A | IHC | Loss atyp. EM | Het loss | |

**Case 4 Detected Alterations**

| Gene | Hotspot | DIE Rectum | OMA L.ovary | EM Cul-de-sac |
|---|---|---|---|---|
| PIK3CA | p.Glu542Lys | X | | X |
| PIK3CA | p.Glu545Lys | X | | X |
| PIK3CA | p.Gly1049Ser | X | | X |
| PIK3CA | p.Met1043Ile | X | X | X |
| NRAS | p.Gly13Asp | X | X | X |
| CTNNB1 | p.His36Tyr | X | X | X |
| CTNNB1 | p.Gly34Glu | X | | X |
| KRAS | p.Gly12Asp | X | X | X |

**Case 8 Detected Alterations**

| Gene | Hotspot | OMA L. ovary | DIE Appendix | EM* R.pel.wall |
|---|---|---|---|---|
| PIK3CA | p.Glu545Lys | X | | |
| NRAS | p.Gly13Asp | X | X | |
| CTNNB1 | p.Ser37Phe | X | X | |
| KRAS | p.Gly12Val | X | | |
| PTEN | IHC | | Het loss | Het loss |

*Insufficient size for sequencing.

**Case 13 Detected Alterations**

| Gene | Hotspot | DIE Bladder | EM R.abd.wall |
|---|---|---|---|
| EGFR | p.Thr790Met | X | X |
| CTNNB1 | p.His36Tyr | | X |
| CTNNB1 | p.Thr41Ile | | X |
| ERBB2 | p.Ser310Phe | | X |

**Detected Alterations in Simple Cases**

**Case 6 Detected Alterations**

| Gene | Hotspot | EM L.pel.wall | DIE R.su.lig. |
|---|---|---|---|
| PIK3CA | p.Met1043Ile | X | X |

**Case 9 Detected Alterations**

| Gene | Hotspot | DIE Appendix | OMA R.ovary |
|---|---|---|---|
| PIK3CA | p.Gln546Arg | | X |
| KRAS | p.Gly12Ala | X | |
| KRAS | p.Gly12Cys | X | |
| KRAS | p.Gly12Val | | X |

**Case 19 Detected Alterations**

| Gene | Hotspot | EM Pararec.Space | DIE Sigmoid |
|---|---|---|---|
| KRAS | p.Gly12Ser | X | X |
| PTEN | IHC | Het loss | |

**Case 20 Detected Alterations**

| Gene | Hotspot | EM Bladder | DIE L.su.lig |
|---|---|---|---|
| KRAS | p.Gly12Asp | | X |
| PTEN | IHC | Loss | |

**Case 21 Detected Alterations**

| Gene | Hotspot | OMA R. ovary | DIE Rv.septum | OMA L. ovary | DIE L.su.lig | DIE Bladder |
|---|---|---|---|---|---|---|
| KRAS | p.Gly12Asp | X | X | | X | X |

**Case 22 Detected Alterations**

| Gene | Hotspot | DIE R.su.lig | EM Cul-de-sac | DIE L.su.lig |
|---|---|---|---|---|
| KRAS | p.Gly12Ala | | X | |

**Case 24 Detected Alterations**

| Gene | Hotspot | DIE Right psoas | DIE Rv. septum | DIE L.su.lig. |
|---|---|---|---|---|
| EGFR | p.Thr790Met | | | X |
| PTEN | IHC | | Het loss | |

**Figure S2:** Per-Case summary of detected alterations across lesions as derived from sequencing, digital droplet PCR (ddPCR), and immunohistochemistry (IHC) in informative cases. Legend: Abd = abdominal, atyp= atypical, DIE= deep infiltrating endometriosis, EM= superficial endometriosis, Het loss= heterogeneous loss, l= left, lig= ligament, pararec=pararectal, OMA= endometrioma, r=right rv= rectovaginal, su= sacrouterine
